# Strain patterns with ultrasound for improved assessment of abdominal aortic aneurysm vessel wall biomechanics

**DOI:** 10.1101/2024.05.27.24307963

**Authors:** Ulver S. Lorenzen, Marta I. Bracco, Alexander H. Zielinski, Magdalena Broda, Stéphane Avril, Laurence Rouet, Jonas P. Eiberg, the COACH Research Collaborative

## Abstract

**Background:** Abdominal aortic aneurysms (AAAs) are an important cause of death. Small AAAs are surveyed with ultrasound (US) until a defined diameter threshold, often triggering a CT scan and surgical repair. Nevertheless, 5-10 % of AAA ruptures are below threshold, and some large AAAs never rupture. AAA wall biomechanics may reveal vessel wall degradation with potential for patient-centred risk assessment. This clinical study investigated AAA vessel wall biomechanics and deformation patterns, including reproducibility.

**Methods:** In 50 patients with AAA, 183 video clips were recorded by two sonographers. Prototype software extracted AAA vessel wall principal strain characteristics and patterns. Functional principal component analysis (FPCA) derived strain pattern statistics.

**Results:** Strain patterns demonstrated reduced AAA wall strains close to the spine. The strain pattern ‘topography’ (i.e., curve phases or ‘peaks’ and ‘valleys’) had a 3.9 times lower variance than simple numeric assessment of strain amplitudes, which allowed for clustering in two groups with FPCA. A high mean reproducibility of these clusters of 87.6 % was found. Median pulse pressure-corrected mean principal strain (PPPS) was 0.038 %/mmHg (interquartile range: 0.029 to 0.051 %/mmHg) with no correlation to AAA size (Spearman’s ρ = 0.02, FDR-P = 0.15). Inter-operator reproducibility of PPPS was poor (limits of agreement: ±0.031 %/mmHg).

**Discussion:** Strain patterns challenge previous numeric stiffness measures based on AP-diameter and are reproducible for clustering. This study’s PPPS aligned with prior findings, although clinical reproducibility was poor. In contrast, US-based strain patterns hold promising potential to enhance AAA risk assessment beyond traditional diameter-based metrics.

## INTRODUCTION

Abdominal aortic aneurysms (AAA) are characterised by progressive vessel wall weakening, with a prevalence of about 2-3 % among 65-year-old men (1, 2). This weakening can lead to rupture with a mortality of 75-80 % (3). In contrast, the mortality associated with prophylactic AAA repair, while comparatively low, remains non-negligible, ranging from 1-3 % (4). Overall, AAA is estimated as the 10^th^-15^th^ most common cause of death among late middle-aged men (5, 6). Repair is typically considered for large AAAs, assessed by diameter, when the risk of rupture exceeds the procedure-related risk. Conversely, small AAAs are routinely monitored with ultrasound (US) surveillance (7–10). However, using diameter as the sole parameter for risk assessment may be flawed since 5-10 % of all AAAs ruptures occur below the threshold, while other AAAs grow large without rupturing (11–16). Declining rupture rates, attributed to AAA screening programs and smoking cessation (2, 17), combined with a potential need for screening more senior citizens and patients with AAA ectasia (18), may place a capacity constraint on AAA surveillance programs. Thus, a more personalised approach is needed to detect subthreshold AAAs at risk and prevent overtreatment of low-risk AAAs above the threshold.

In search of AAA risk factors beyond diameter, biomechanical properties, especially global stiffness and elastic modulus of the AAA vessel wall, have been studied with US. Both are typically calculated based on the anterior-posterior (AP) diameter dilatation within a cardiac cycle, normalised for blood pressure (19–24). These studies correlated the global stiffness and elastic modulus with AAA diameter, and the results ranged from negligible (19, 24) to moderate correlation (21, 23). Similarly, studies have investigated the correlation between biomechanical properties and future growth (23, 24), rupture, or repair (20–22). Despite initially promising results, the evidence remains contradictory and does not consistently align with findings from in vitro studies (25).

Further work has focused on localised AAA wall strains, calculated by mapping the AAA wall motion with time-resolved two-dimensional (2D-US) (26, 27) and three-dimensional US (3D-US) (28–32). Local strains are defined by deformation of hypothetical small segments distributed throughout the AAA vessel wall. This allows for the computation of an entire grid of local AAA vessel wall deformation across the cardiac cycle, or a “strain map”. One study found that such strain measures are independent of AAA size and have a non-linear correlation to AAA growth, suggesting that localised AAA wall strains can potentially improve the current diameter-based AAA risk prediction (27).

Thus, progress has been made from simple global stiffness and elastic modulus solely based on ultrasound AP-diameter variations to full strain maps. However, only one study (27) has used these new biomechanical markers in AAA risk prediction and did not investigate clinical reproducibility. Also, all current studies of localised wall strains reduce the full AAA vessel wall deformation grid, or strain map, to “single parameter characterisations”. Such characterisations include numeric mean strain, peak strain, and heterogeneity index, which all can be calculated from the strain values of the entire grid. These numeric characterisations, while ingenious, can hide nuances that can only be obtained by considering the complete ‘topography’ of a full AAA strain map.

This paper aimed to measure and categorise localised AAA strain maps in a clinical setting using outpatient-based 2D-US acquisitions and a prototype software tool based on a fast strain-mapping approach and to investigate the method’s reproducibility.

## MATERIALS & METHODS

### Study design

A prospective single-centre, observational proof-of-concept study

### Patients

Fifty patients from the Copenhagen Aortic CoHort (COACH) were included. COACH is a prospective cohort of patients with AAAs with diameters above 30 mm and monitored with research-orientated US surveillance (33). Enrolment in COACH relies on research staff and equipment availability. Exclusion criteria were aorto-iliac aneurysms; suprarenal aortic aneurysms; prior aortic or iliac artery repair; non-compliance with surveillance; or inability to provide informed consent. US acquisitions were systematically collected and stored.

## METHODS

### US image acquisition

Two US operators independently and non-consecutively recorded two 10-second cross-sectional 2D-US time-resolved B-mode sequences (video clips called “cineloops”) of the AAA at the maximal AP-diameter during breath-holds using an identical US-system (Epiq Elite US system and C5-1 transducer, Philips Medical systems, Bothell, WA, USA). Thus, for each patient, four cineloops were planned (two recordings × two operators). Acquisitions were subsequently transferred to a core lab, where two readers (USL and MIB) mutually and independently conducted the image post-processing in a randomised order via randomised.org, using a prototype software (Philips Health Technology innovation). The image quality of every cineloop was evaluated and graded by a clinical expert reader (USL). Cineloops of poor image quality were not used for further analysis.

### Strain

Strains are the percentage deformation of the AAA vessel wall when force is applied to it, e.g., by the blood pressure during the heart cycle. When overlaying a grid on the AAA vessel wall, principal strain is the largest deformation value in any direction (called the “principal direction”). Higher values indicate softer tissues, and lower values indicate stiffer tissues. Principal strain can then be projected onto two main directions of movement along the AAA vessel wall: circumferential (stretch or compression) or radial (thickening or thinning); see Figure S1.

### Image post-processing

The image post-processing is described in detail in previous work (26), with the notable difference that the present study includes principal strain in addition to circumferential and radial strain. The process can be summarised as a 3-step procedure: 1) manual segmentation and automatic tracking; 2) automatic strain map generation; and 3) strain pattern extraction.

#### 1) Manual segmentation and automatic tracking

Tracking points were manually positioned at the initial frame of the cineloop. This was done circumferentially at the interface between the aneurysm wall and lumen (Figure 1B). Subsequently, an automated speckle-tracking process was employed based on image gradients within an automatic region of interest (Figure 1C).

**Figure 1:**
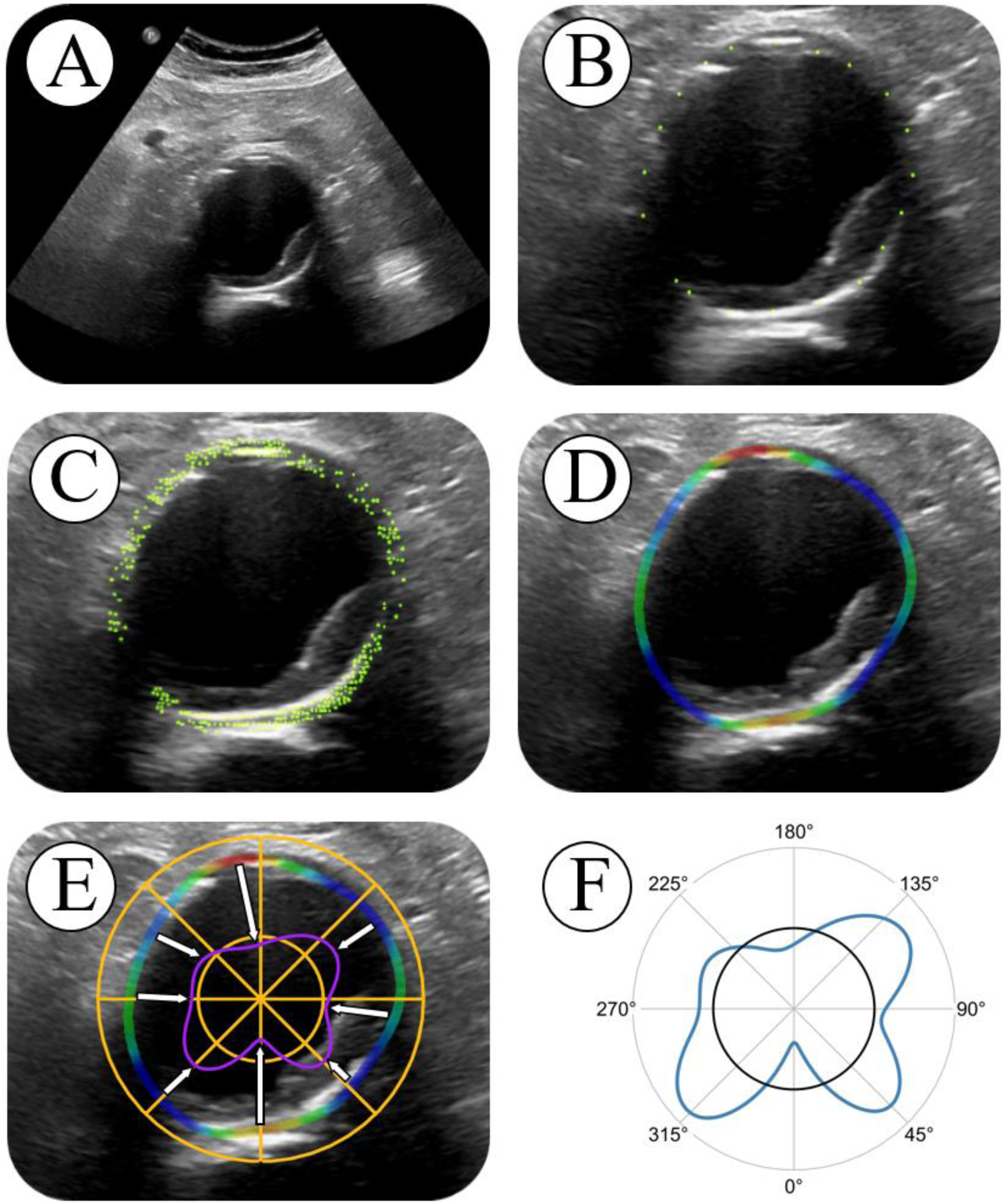
Overview of procedure for strain pattern generation. The steps are: A) acquisition of a 10-second cineloop; B) manual segmentation, placing tracking points along the AAA wall/lumen border; C) automatic speckle tracking; D) generation of strain map; E) extrapolation of strain values to a circle; and F) plotting the strain pattern in a polar plot.

#### 2) Automatic strain map generation

A deformation grid was computed based on the tracking procedure, and from this, strain over the cardiac cycle – from end diastole to peak systole – was directly calculated. The resulting strain data were superimposed on the cineloop. This generated a comprehensive strain map (Figure 1D) illustrating different zones of AAA strain, expressed as percentage changes over time. Results from all cardiac cycles were then averaged.

#### 3) Strain pattern extraction and normalisation

The obtained strain maps contain local information along an irregular vessel wall. To enable inter-case comparisons of strain maps, strain values were radially averaged for each degree of the AAA circumference with 0° defined as the bottom of the strain map (towards the spine), see Figure 1E. Thus, the total information of an AAA strain map is summarised as a polar plot, henceforth designated the “strain pattern”, see Figure 1F (where the black circle represents the reference null strain). Strain patterns were then smoothed to reduce sensitivity to noise.

### Data analysis and statistics

#### Functional data analysis of strain patterns

To analyse strain patterns without reducing them to single parameter characterisations, functional data analysis (FDA) with alignment was performed (34). Alignment is the process of elastically synchronizing (or “warping”) functional data sets to a common reference, enabling comparative analysis and inference across patients. The mean strain pattern was calculated on smoothed and elastically aligned strain patterns to reduce noise and account for minor scan variations (e.g., in insonation angles). Further, using function principal component analysis (FPCA), the strain patterns were decomposed into their principal modes of variations (PMV) with associated scores for each cineloop. The scores for each PMV was analysed for correlation with AAA size using Spearman’s rank correlation. A positive or negative coefficient of 0 to 0.3 was considered weak correlation, 0.3 to 0.5 low, 0.5 to 0.7 moderate, 0.7 to 0.9 high, and 0.9 to 1.0 very high (35). P-values were corrected using the Benjamini-Hochberg false discovery rate (FDR) procedure. Only FDR-P-values < 0.05 were considered statistically significant.

#### Clustering

Subsequently, spectral consensus clustering (36, 37) was then used to split the cineloops into an optimal number of clusters based on the PMV scores.

#### Single parameter strain characterisations

As described in other work on strain mapping (27–29), single numeric parameter strain characterisations can be calculated from the end-diastolic to peak-systolic strain based on the principal strains across the vessel wall: mean strain; pulse pressure-normalised mean strain (PPPS); peak strain; heterogeneity index, ie., the ratio of the SD of wall strain to the mean wall strain; and local strain ratio, i.e., the ratio between the peak strain and mean strain.

Single parameter strain characteristics were summarised across all patients with frequency-weighted medians and interquartile ranges (IQR), due to non-normality. All single parameter strain characteristics were analysed for correlation with AAA size using Spearman’s rank correlation similarly to the PMVs.

#### Reproducibility

For strain pattern clusters, the “cluster reproducibility” was calculated, which was defined as the percentage of an individual patient’s four cineloops that belong to the majority (or tie for majority) cluster (e.g., three out of four scans within the same cluster equals 75 %). By aligning all of a patient’s scans to each other (within-patient alignment), amplitude and phase variations for each patient could be extracted. These were averaged across patients for an overall amplitude-phase variation ratio.

For single parameter characterisations, Bland-Altman limits of agreement (LoA) on inter-reader comparisons were calculated, comparing readers USL and MIB on the same cineloops. The LoA were quantile-based, due to non-normality, and were converted into a range of variance (RoV, i.e. half the quantile distance) and displayed as a percentage of the median. To calculate LoA for inter-operator considerations, a linear mixed-effects model was utilised, accounting for multiple acquisitions by the same operator.

All statistical analyses described above were performed in R version 4.3.3 using RStudio version 2023.12.1 build 402 (Posit software, PBC, Boston MA, USA). The packages: Hmisc, mgcv, fdasrvf, fdapace, and M3C were used for weighted statistics, strain pattern smoothing, alignment, FPCA, and clustering, respectively.

### Ethics

This study was approved by the The Regional Ethics Committee of the Capital Region of Denmark (protocol no. H-20001116).

## RESULTS

The present study utilised a prototype US tool to estimate AAA vessel wall strains from 183 clinical US cineloops acquired on 50 AAA patients scanned by two independent operators. The frequency-weighted median AAA size with US was 45.1 mm (weighted IQR: 41.7 to 49.4 mm), and all AAA’s were asymptomatic. Further demographics can be seen in Table 1.

**Table 1:** Demographics table. SD = standard deviation. N = patients with demographics data. NYHA = New York Heart Association score.

|  | Overall (N=47) |
| --- | --- |
| **Sex** |  |
| Female | 9 (19 %) |
| Male | 38 (81 %) |
| **Age (years)** |  |
| Mean (SD) | 73 (4.7) |
| **Height (m)\*** |  |
| Mean (SD) | 1.8 (0.075) |
| **Weight (kg)\*** |  |
| Mean (SD) | 85 (17) |
| **Body mass index (kg/m<sup>2</sup>)\*** |  |
| Mean (SD) | 27 (4.7) |
| **Hypertension** |  |
| No | 14 (30 %) |
| Yes | 33 (70 %) |
| **Ischaemic heart disease** |  |
| No | 40 (85 %) |
| Yes | 7 (15 %) |
| **NYHA** |  |
| None | 5 (11 %) |
| I | 39 (83 %) |
| II | 2 (4 %) |
| III | 1 (2 %) |
| IV | 0 (0 %) |
| **Blood thinner** |  |
| None | 7 (15 %) |
| Platelet inhibitor | 36 (77 %) |
| Anticoagulant | 3 (6 %) |
| Dual-pathway inhibition | 1 (2 %) |
| **Cholesterol lowering** |  |
| No | 8 (17 %) |
| Yes | 39 (83 %) |
| **Smoking\*** |  |
| Never | 2 (4 %) |
| Past | 24 (51 %) |
| Current | 20 (43 %) |
| **Diabetes\*** |  |
| No | 37 (79 %) |
| Yes | 9 (19 %) |
| **Pulmonary disease** |  |
| None | 39 (83 %) |
| Obstructive | 7 (15 %) |
| Restrictive | 1 (2 %) |

\*Further n = 1 missing.

### Strain patterns

The strain patterns were extracted as the strain values at each degree on a circle and illustrated as polar plots in Figure 2. The mean principal strain pattern behaved as a triphasic, though almost quadriphasic, curve with three definitive local maxima and a plateau. At the posterior wall, two definitive peaks (3.5 % and 3.2 %) were detected on each side of the spine (320° and 60°). At the anterior wall, one definitive peak (2.1 % at 244°) was identified at the patient’s right. In order words, AAA principal strain patterns reveal that, on average, the vessel wall was the softest bilaterally adjacent to the spine and on the right side of the anterior wall while demonstrating maximal stiffness directly over the spine. The mean strain patterns were generally consistent across the two operators. Results for circumferential and radial strain can be seen in Figure S2 and Table S1.

**Figure 2:**
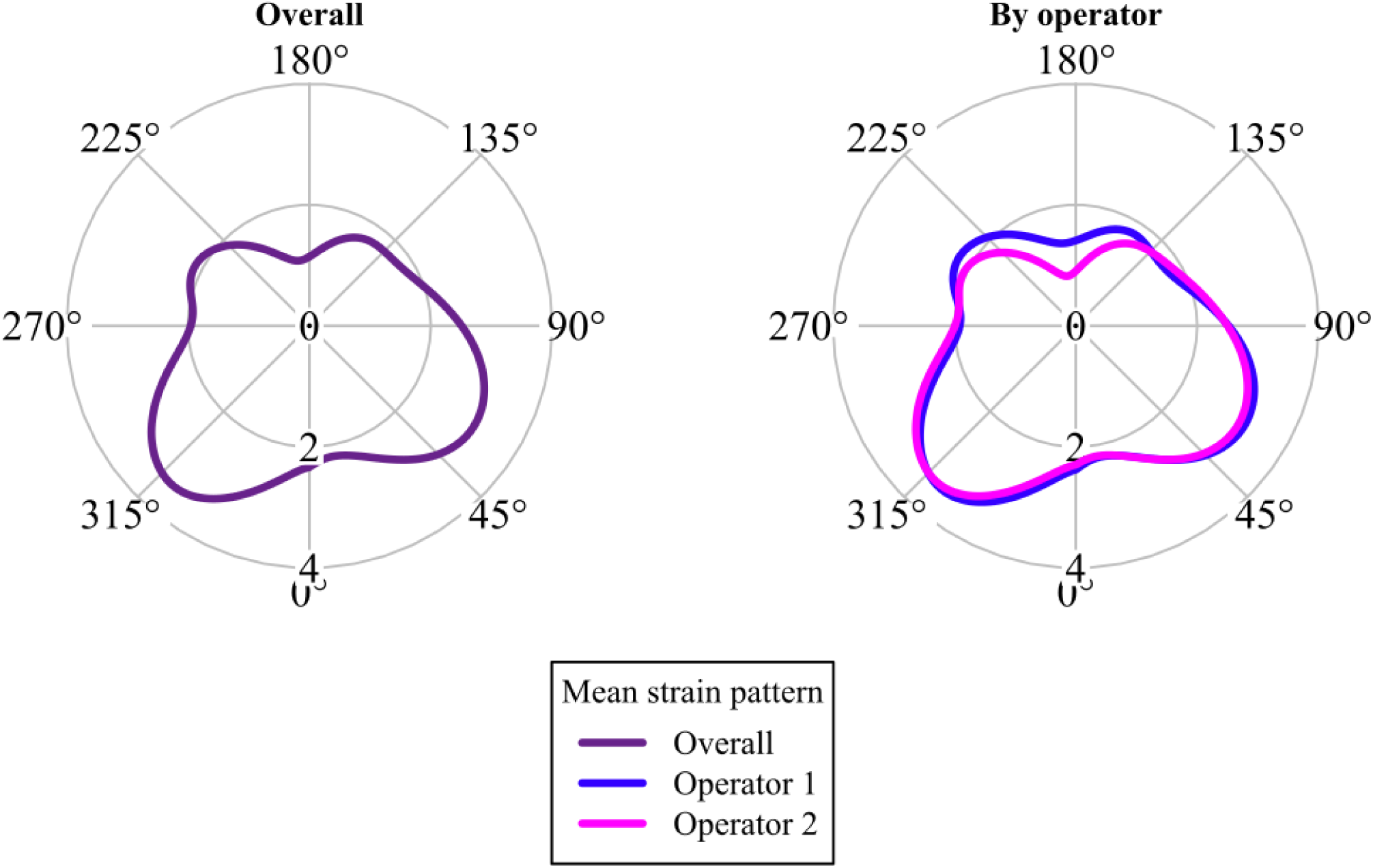
Polar plots illustrating aligned mean principal strain patterns across all cineloops from all patients. Larger values indicate softer tissues and smaller values indicate stiffer tissues. The averages are shown comparing operators.

Three of the eight PMVs extracted with FPCA (see Figure S3) showed evidence of a statistically significant correlation with AAA size (Table 2). The PMV#2 had a ρ of −0.24 (FDR-P = 0.005), PMV#4 had a ρ of −0.20 (FDR-P = 0.02), and PMV#8 had a ρ of 0.29 (FDR-P < 0.001). The PMVs with the most negative and positive correlations to AAA size, PMV#2 and #8, are illustrated in Figure 3 alongside scatterplots of their correlations to AAA size. From this, it can be inferred that large AAAs tend towards more strain at the anterior wall with two peaks to the right and left side, while small AAAs tend towards less strain at the anterior wall, but retaining strain peaks in relation to the spine. Also, these two PMVs show that large AAAs tend to have less strain at the lateral walls, while small have more.

**Figure 3:**
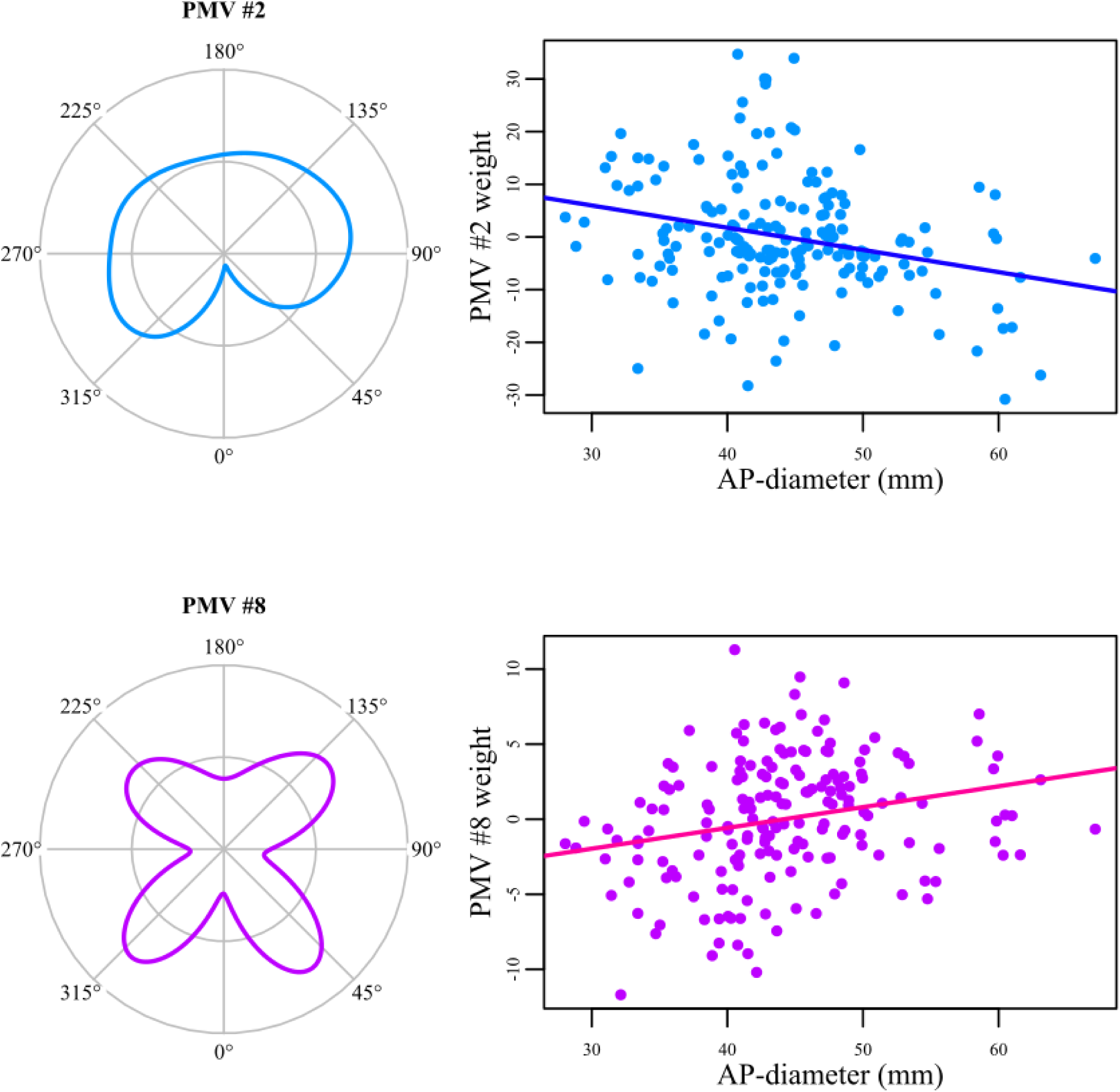
Polar plots of the principal mode of variation (PMV) with the most negative correlation (PMV #2) and of the PMV with the most positive correlation (PMV #8) to abdominal aortic aneurysm (AAA) size. These are placed next to a scatterplot of the PMV score as a function of AAA size with a simple linear trend line for clarity.

**Table 2:** Correlations between abdominal aortic aneurysm (AAA) size and each of the principal strain pattern principal modes of variation (PMV) from the functional principal component analysis. ρ = Spearmans’ ρ, P = P-value, FDR-P = false discovery rate-corrected P-value. Statistically significant FDR-P is marked with *.

| PMV | Correlation to AAA size |  |  |
| --- | --- | --- | --- |
| | $\rho$ | P | FDR-P |
| #1 | -0.02 | 0.78 | 0.78 |
| #2 | -0.24 | 0.001 | 0.005* |
| #3 | 0.13 | 0.07 | 0.15 |
| #4 | -0.20 | 0.008 | 0.02* |
| #5 | -0.09 | 0.20 | 0.33 |
| #6 | 0.07 | 0.35 | 0.46 |
| #7 | -0.05 | 0.54 | 0.62 |
| #8 | 0.29 | <0.001 | <0.001* |

### Clustering

With spectral consensus clustering, the strain patterns from all cineloops were split into an optimum of two clusters by the algorithm (P = 0.04), the aligned means of which can be seen in Figure 4 (and Figure S4). Generally, the clusters’ aligned mean strain patterns were similar in shape, but differed in the amplitudes of the shape.

**Figure 4:**
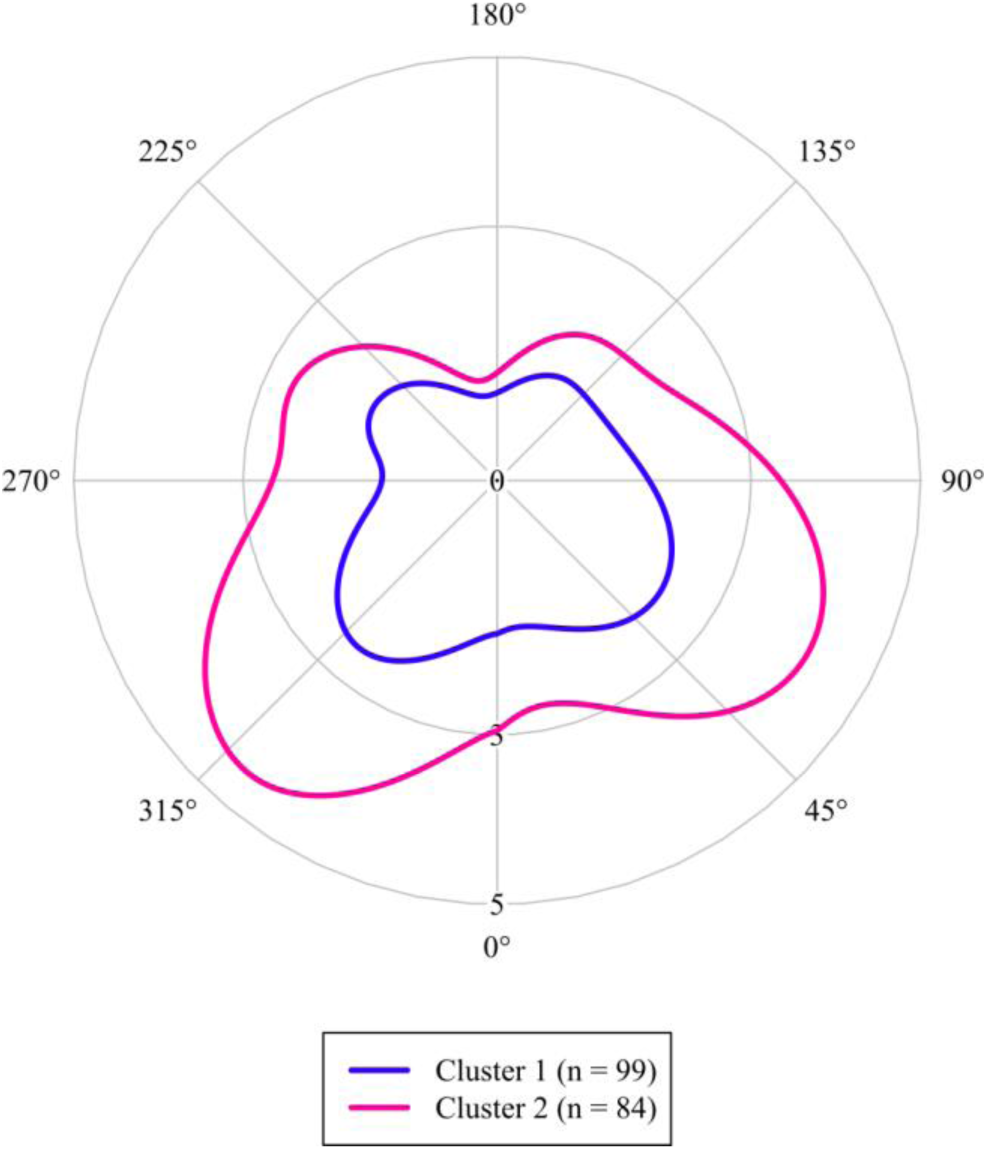
The mean principal strain patterns split into two clusters by spectral consensus clustering. Note scale difference compared to Figure 2.

### Single parameter characterisations

Single parameter characterisations are summarised in Table 3. The pulse pressure-corrected mean principal strain (PPPS) displayed a weighted median of 0.038 %/mmHg (IQR: 0.029 to 0.051 %/mmHg). As also seen in Table 3, none of the single parameter characterisations showed evidence of correlation with AAA size, indicating that these values contain additional information to AAA diameter.

**Table 3:** Frequency-weighted medians and interquartile ranges (IQR) along with Spearman’s rank correlation with abdominal aortic aneurysm (AAA) size for each of the principal strain single parameter characterisations. PPPS = pulse pressure-normalised mean principal strain, ρ = Spearmans’ ρ, P = P-value, FDR-P = false discovery rate-corrected P-value.

| Parameter | Overview |  | Correlation to AAA size |  |  |
| --- | --- | --- | --- | --- | --- |
| | Median | IQR | $\rho$ | P | FDR-P |
| Mean strain (%) | 2.1 | (1.7 to 2.6) | -0.02 | 0.80 | 0.94 |
| PPPS (%/mmHg) | 0.038 | (0.029 to 0.051) | 0.09 | 0.22 | 0.94 |
| Peak strain (%) | 4.6 | (3.4 to 6.1) | 0.01 | 0.94 | 0.94 |
| Heterogeneity index | 0.54 | (0.44 to 0.65) | 0.02 | 0.76 | 0.94 |
| Local strain ratio | 2.1 | (1.8 to 2.4) | 0.06 | 0.41 | 0.94 |

### Reproducibility

Mean cluster reproducibility across patients were 86.2 % (95 % CI: 81.4 % to 91.1 %) for principal strain (range of possible values with two clusters and four scans pr. patient is 50-100 %, see also Figure S5). From the alignment procedure, amplitude and phase variations were extracted. Amplitude-phase variance ratios were calculated for principal strain and was 3.9 (95 % CI: 3.3 to 4.7) on average. Thus, the phases of the strain patterns were vastly more reproducible than their specific amplitudes. Example cases with high amplitude-phase variance ratios can be seen in Figure S6.

Bland-Altman statistics comparing two readers post-processing the same images were converted to RoVs. The PPPS’ inter-reader RoV was low at 0.007 %/mmHg, which translated to 20 % of the median value, indicating good reproducibility of the post-processing. The RoVs for other strain parameters were similar, ranging from 14 % for local strain ratio to 22 % for peak strain. Based on a linear mixed-effects model, inter-operator LoA were calculated; for PPPS this was quite high at ±0.031 %/mmHg (82 % of the median). Further reproducibility of principal strain single parameter characterisations can be seen in Table 4. The principal strain was the most reproducible method compared to circumferential and radial strain in Table S3.

**Table 4:** Inter-reader and inter-operator reproducibility for principal strain single parameter characterisations, denoted by range of variance (RoV) and limits of agreement (LoA) from the quantile-based Bland-Altman statistics and the linear mixed-effects model, respectively. Both RoV and LoA are shown as absolute values (percentage of median).

| Parameters | Inter-reader | Inter-operator |
| --- | --- | --- |
|  | RoV | LoA |
| Mean strain (%) | 0.33 (16 %) | 1.4 (69 %) |
| PPPS (%/mmHg) | 0.007 (20 %) | 0.031 (82 %) |
| Peak strain (%) | 0.99 (22 %) | 3.7 (81 %) |
| Heterogeneity index | 0.099 (19 %) | 0.43 (82 %) |
| Local strain ratio | 0.29 (14 %) | 1.2 (55 %) |

## DISCUSSION

Vessel wall biomechanics have been explored to discover future AAA risk factors. Global stiffness and elastic modulus based on AP-diameter variations seem inconclusive (20, 22, 24), and this study presents an approach using conventional 2D-US to detect local strain changes along the entire AAA circumference.

When measuring the strain on 183 cineloops in 50 patients, notable variations in the localised strain were detected, indicating that ‘global stiffness’ and elastic modulus, based on AP-diameter variations, seem oversimplified. These ‘topographic’ variations were closely related to the spine, suggesting that the surrounding tissue has a profound, and perhaps overlooked, impact on AAA wall biomechanics. For ease of interpretation, earlier works have reduced these localised variations (called ‘strain patterns’ in the present paper) into numeric parameters as a simplified description of the AAA vessel wall motion (26–32). Of these, one (27) has investigated strain mapping from conventional 2D-US, and have reduced the strain patterns to pulse pressure-corrected mean principal strains (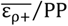, here called PPPS), finding evidence of a connection between intermediate values at baseline and future AAA growth. Interestingly, they found a mean PPPS of 0.034 %/mmHg in 113 AAA patients, which is very close to the weighted median in the present study (0.038 %/mmHg). This similarity in results implies that the measured phenomenon reflects genuine deformation characteristics of the AAA vessel wall.

From the current work on strain patterns, the shape, or ‘topography’, of the AAA wall motion curve is quite reliable and easy to reproduce across operators. An example is the almost four-fold decrease in variance when looking at strain pattern phases rather than amplitudes. This is particularly important because, even though a few measures (e.g. heterogeneity index) superfluously describe characteristics of local variations, all characteristics described in earlier work entirely ignore positions of strain variations along the AAA vessel wall. When considering these shapes, clustering based on strain patterns becomes feasible and reproducible, as illustrated by the 86 % cluster reproducibility for principal strain, meaning that an average of about 3.5 out of 4 cineloops belong to the same cluster. Strain pattern clusters are a data-driven way to stratify AAAs. The clusters may reflect important differences in wall characteristics; in this initial study, about half of the patients exhibited stiffer aneurysms. Theoretically, these clusters could be correlated with AAA growth and even rupture.

Compared to the reproducibility of strain pattern clusters, Zottola et al. (27) investigated the reproducibility of US-generated AAA vessel wall motion characteristics. On the log scale, they reported an LoA of PPPS that can be translated into an RoV of 15.6 % – slightly better than the RoV of 20 % found in the present study. A notable difference between these methods, is that this study averages strains across all cycles of a 10-second cineloop, while Zottola et al. selected a single cycle from each cineloop. Both Zottola and this study reported inter-reader reproducibility. However, the present study is the first to include inter-operator reproducibility, which is essential if the method should play a role in risk assessment in future AAA surveillance. Unfortunately, reproducibility was challenging for all the investigated parameters, with measurement errors as large as the actually measured values. While this does not invalidate correlation and follow-up studies’ ability to detect essential associations, the transfer into clinical practice on an individual patient level becomes problematic. The reproducibility issue may have plagued early studies based on AP-diameter variations (19–24).

Many factors can affect strain measurements, and reproducibility issues were expected. Common ultrasound challenges such as adipose tissue, patient breathing, and bowel movements may additionally have an impact. Strain measurement can theoretically be further hampered by operator-related issues such as insonation angle, cross-section deviations, transducer rotation, and even transducer pressure (38–40). Unlike in diameter measurement, patient factors such as heart rate and blood pressure may also limit strain measurements. Despite this, strain patterns appear markedly stable, and their relation to AAA size suggests that AAA strain patterns should be investigated as a viable supplement for a more patient-centred AAA risk stratification. It is conceivable that the progression and rupture of AAA are influenced not by the overall AAA strain but more by localised vessel wall weakness, or ‘weak spots’, and the dynamic interactions with adjacent tissue structures. The surrounding tissue must be essential in restricting AAA wall motion (38), which should also be considered.

### Limitations

Further studies with follow-up data should determine the added clinical value of strain pattern analysis and AAA biomechanics for improved AAA risk prediction. Larger cross-sectional studies could provide insights into a correlation between biomechanical measures and patient characteristics: demographics, diabetes, hypertension, medication and sex.

The use of repeated measurements mitigates the impact of the relatively small size of this study; however, because of repeated measures and FDA, a preceding power calculation was unfeasible. Therefore, these results must be evaluated considering a potential Type II error.

### Current and future work

With the reservation that this is not a follow-up study, this study suggests that, as the AAAs grow, strain pattern changes may occur at the anterior AAA wall, while the posterior wall strain is constant. The lateral wall strains also showed relation to AAA diameter, though these locations are subject to poorer image quality. Further investigation into these correlations will be an important feature in future studies.

### Conclusion

This proof-of-concept study presents the first experiences for detecting, visualising, and quantifying circumferential AAA vessel wall strain patterns based on conventional US scanning. Interestingly, deformation along the circumference of the aneurysm wall was heterogenous, specifically for the wall close to the spine. These results open the possibility for prospective studies investigating not just global, but local strain as a risk factor for growth.

## Data Availability

All data produced in the present study are available upon reasonable request to the authors

## ACKNOWLEDGEMENTS

Members of the Copenhagen Aneurysms Cohort (COACH) Research Collaborative that are not authors on this paper, but who collected data and/or developed the AAA tools, and/or analysed and interpreted data, and/or developed study ideas and design:

Cecile Dufour; Qasam M. Ghulam

This study did not obtain specific funding from public, commercial, or not-for-profit sectors.

## CONFLICTS OF INTEREST

**Jonas Eiberg** has received a research grant, speaker honorarium and sits on an advisory board for Philips Ultrasound.

**Laurence Rouet** is currently employed at Philips^®^ Ultrasound.

**Marta Bracco** was employed at Philips^®^ Ultrasound.

**Stéphane Avril** is currently employed ***

**Ulver Lorenzen**, **Alexander Zielinski**, and **Magdalena Broda** have no relevant conflicts of interest.

## SUPPLEMENTARY MATERIALS

**Table S1:** Local extrema describing the aligned mean curves for the principal, circumferential, and radial strains.

| Principal strain |  | Circumferential strain |  | Radial strain |  |
| --- | --- | --- | --- | --- | --- |
| Position (°) | Strain (%) | Position (°) | Strain (%) | Position (°) | Strain (%) |
| 12 | Min: 2.2 | 9 | Min: -0.9 | 9 | Max: 0.3 |
| 60 | Max: 3.2 | 70 | Max: 2.1 | 55 | Min: -0.6 |
| 189 | Min: 1.1 | 128 | Min: 0.5 | 104 | Max: 0.3 |
| 244 | Max: 2.1 | 152 | Max: 0.7 | 177 | Min: -1.3 |
| 266 | Min: 1.9 | 191 | Min: 0.0 | 274 | Max: 0.4 |
| 320 | Max: 3.5 | 238 | Max: 1.3 | 321 | Min: -0.9 |
|  |  | 271 | Min: 0.4 |  |  |
|  |  | 315 | Max: 2.5 |  |  |

**Table S2:** Correlations between abdominal aortic aneurysm (AAA) size and each of the circumferential and radial strain pattern principal modes of variation (PMV) from the functional principal component analysis. ρ = Spearmans’ ρ, P = P-value, FDR-P = false discovery rate-corrected P-value. Statistically significant FDR-P is marked with *.

| Correlation to AAA size |  |  |  |
| --- | --- | --- | --- |
| PMV | $\rho$ | P | FDR-P |
| Circumferential strain |  |  |  |
| #1 | -0.15 | 0.05 | 0.07 |
| #2 | 0.21 | 0.004 | 0.01* |
| #3 | -0.12 | 0.11 | 0.14 |
| #4 | 0.19 | 0.01 | 0.03* |
| #5 | -0.11 | 0.12 | 0.14 |
| #6 | 0.05 | 0.50 | 0.50 |
| #7 | 0.23 | 0.002 | 0.01* |
| #8 | 0.16 | 0.03 | 0.06 |
| Radial strain |  |  |  |
| #1 | -0.03 | 0.64 | 0.64 |
| #2 | 0.07 | 0.36 | 0.48 |
| #3 | 0.06 | 0.40 | 0.48 |
| #4 | 0.24 | 0.001 | 0.007* |
| #5 | 0.10 | 0.18 | 0.36 |
| #6 | 0.10 | 0.17 | 0.36 |

**Table S3:**
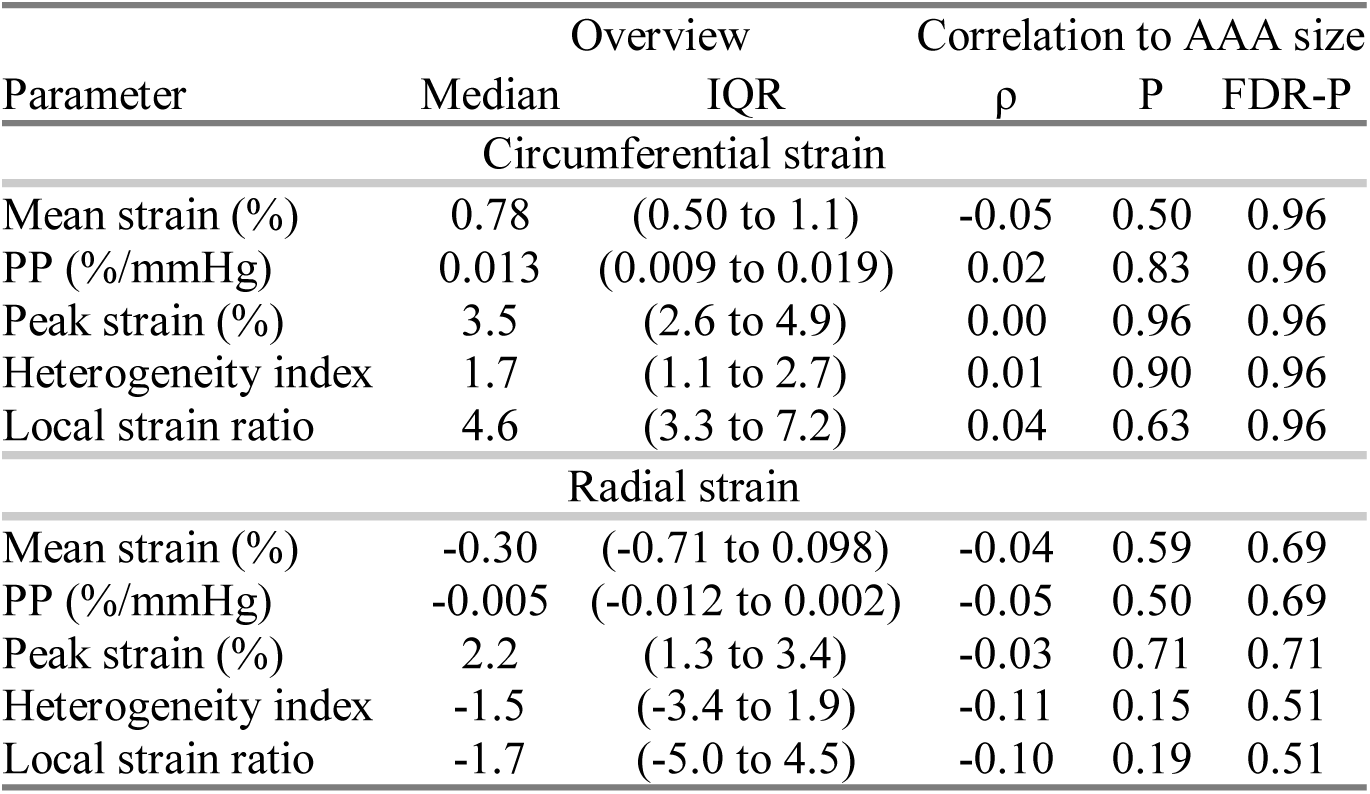
Frequency-weighted medians and interquartile ranges (IQR) along with Spearman’s rank correlation with abdominal aortic aneurysm (AAA) size for each of the principal strain single parameter characterisations. The pulse pressure-normalised mean strains are denoted ‘PP’ and are similar to the pulse pressure-corrected mean principal strains, PPPS, in the paper. ρ = Spearmans’ ρ, P = P-value, FDR-P = false discovery rate-corrected P-value (FDR-P).

| Parameter | Overview |  | Correlation to AAA size |  |  |
| --- | --- | --- | --- | --- | --- |
| | Median | IQR | $\rho$ | P | FDR-P |
| Circumferential strain |  |  |  |  |  |
| Mean strain (%) | 0.78 | (0.50 to 1.1) | -0.05 | 0.50 | 0.96 |
| PP (%/mmHg) | 0.013 | (0.009 to 0.019) | 0.02 | 0.83 | 0.96 |
| Peak strain (%) | 3.5 | (2.6 to 4.9) | 0.00 | 0.96 | 0.96 |
| Heterogeneity index | 1.7 | (1.1 to 2.7) | 0.01 | 0.90 | 0.96 |
| Local strain ratio | 4.6 | (3.3 to 7.2) | 0.04 | 0.63 | 0.96 |
| Radial strain |  |  |  |  |  |
| Mean strain (%) | -0.30 | (-0.71 to 0.098) | -0.04 | 0.59 | 0.69 |
| PP (%/mmHg) | -0.005 | (-0.012 to 0.002) | -0.05 | 0.50 | 0.69 |
| Peak strain (%) | 2.2 | (1.3 to 3.4) | -0.03 | 0.71 | 0.71 |
| Heterogeneity index | -1.5 | (-3.4 to 1.9) | -0.11 | 0.15 | 0.51 |
| Local strain ratio | -1.7 | (-5.0 to 4.5) | -0.10 | 0.19 | 0.51 |

**Table S4:** Inter-reader and inter-operator reproducibility for circumferential and radial strain single parameter characterisations, denoted by range of variance (RoV) and limits of agreement (LoA) from the quantile-based Bland-Altman statistics and the linear mixed-effects model, respectively. Both RoV and LoA are shown as absolute values (percentage of median).

| Parameters | Inter-reader | Inter-operator |
| --- | --- | --- |
|  | RoV | LoA |
| Circumferential strain |  |  |
| Mean strain (%) | 0.20 (26 %) | 1.0 (128 %) |
| PP (%/mmHg) | 0.004 (29 %) | 0.021 (152 %) |
| Peak strain (%) | 0.92 (27 %) | 3.4 (97 %) |
| Heterogeneity index | 4.1 (226 %) | 13 (703 %) |
| Local strain ratio | 8.3 (182 %) | 23 (503 %) |
| Radial strain |  |  |
| Mean strain (%) | 0.40 (129 %) | 1.9 (621 %) |
| PP (%/mmHg) | 0.008 (149 %) | 0.038 (711 %) |
| Peak strain (%) | 1.1 (48 %) | 3.5 (152 %) |
| Heterogeneity index | 42 (2663 %) | 45 (2861 %) |
| Local strain ratio | 72 (4102 %) | 81 (4650 %) |

**Figure S1:**
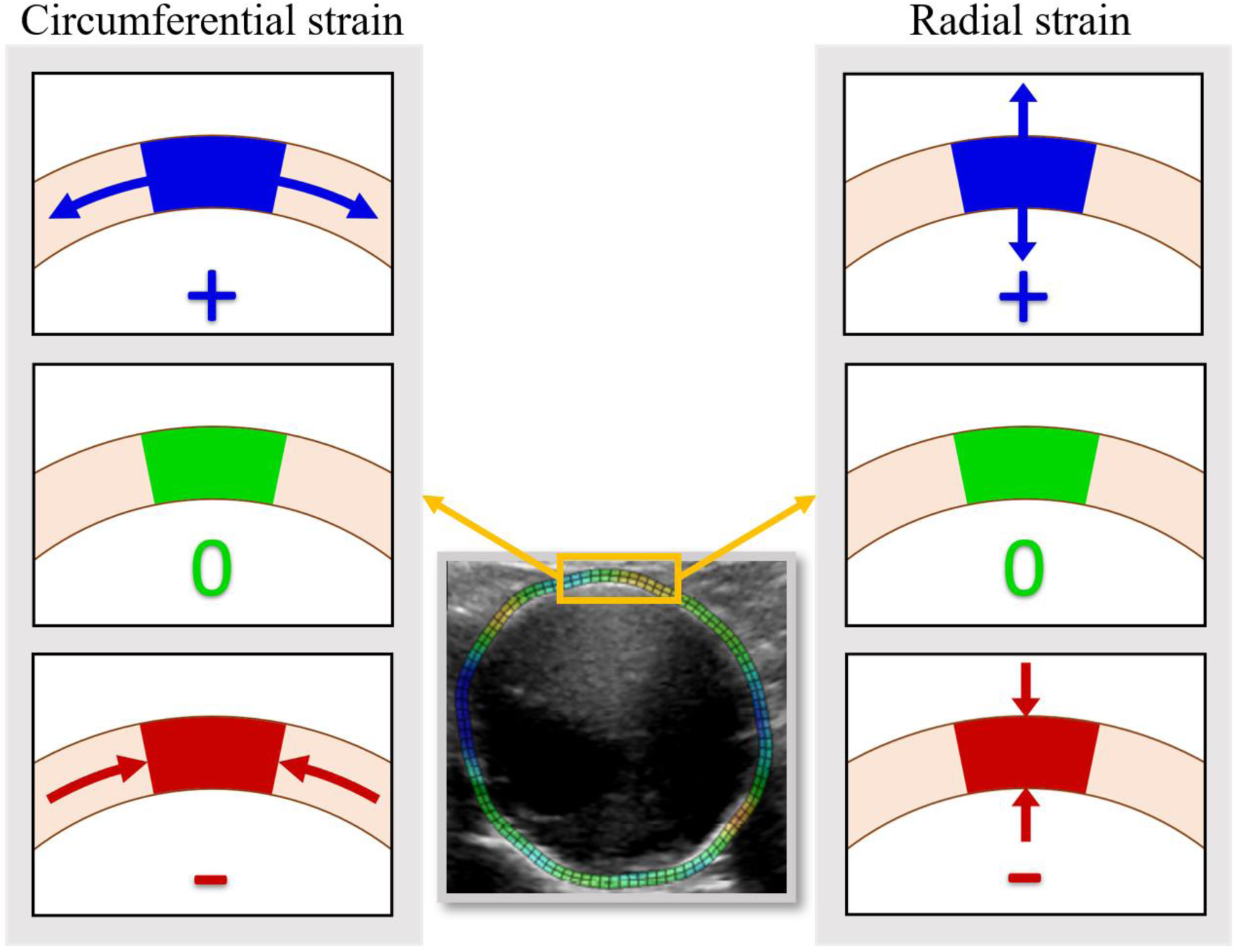
Overview of circumferential and radial strain types with a sample strain map in the centre. Circumferential strain is defined as compression or stretch along the circumference of the AAA vessel wall. Radial strain is thinning or thickening of the vessel wall and surrounding tissue. Both types of strain are visualised in the same manner: zero strain is green, positive strain (stretch) is blue, and negative strain (compression) is red.

**Figure S2:**
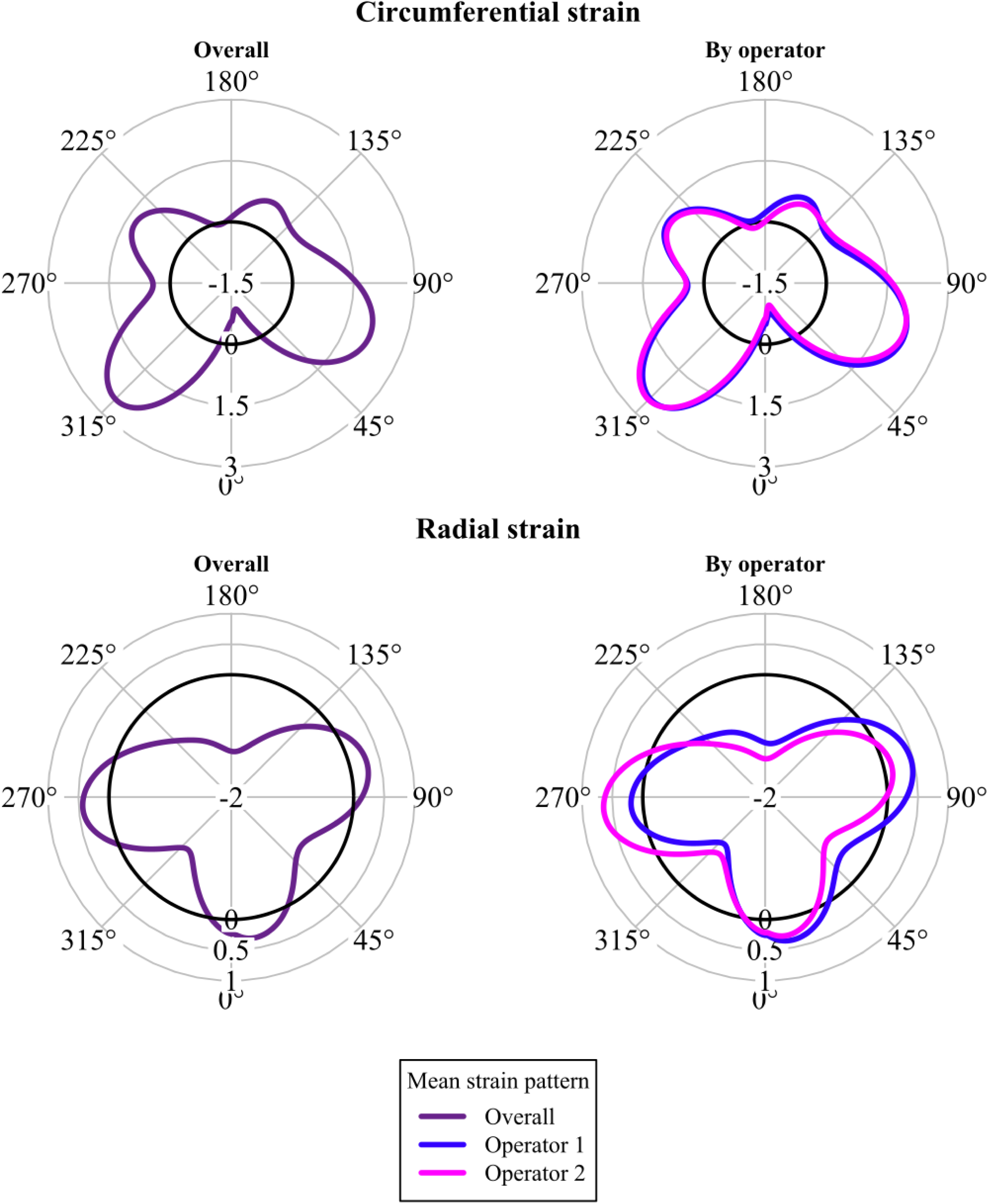
Polar plots illustrating mean circumferential and radial strain across all cineloops. For circumferential and radial strain, respectively, positive values indicate stretching or thickening of the AAA vessel wall, and negative values indicate compression or thinning. The averages are shown comparing operators.

**Figure S3:**
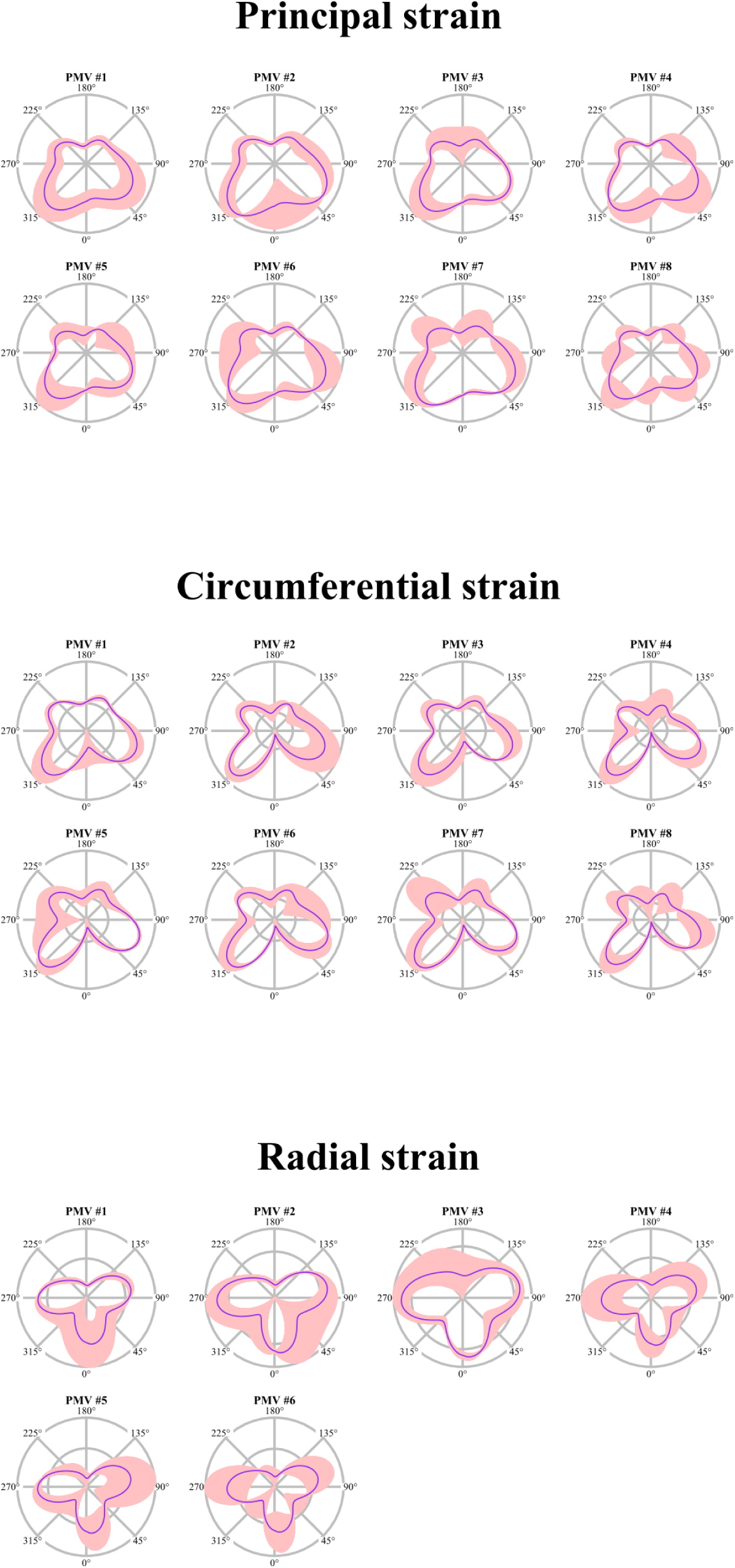
Polar plots of the all principal modes of variation (PMVs) derived from the functional principal component analysis of principal, circumferential, and radial strain. The purple lines show the mean strain pattern, and the pink areas show the mean plus 10 · PMV.

**Figure S4:**
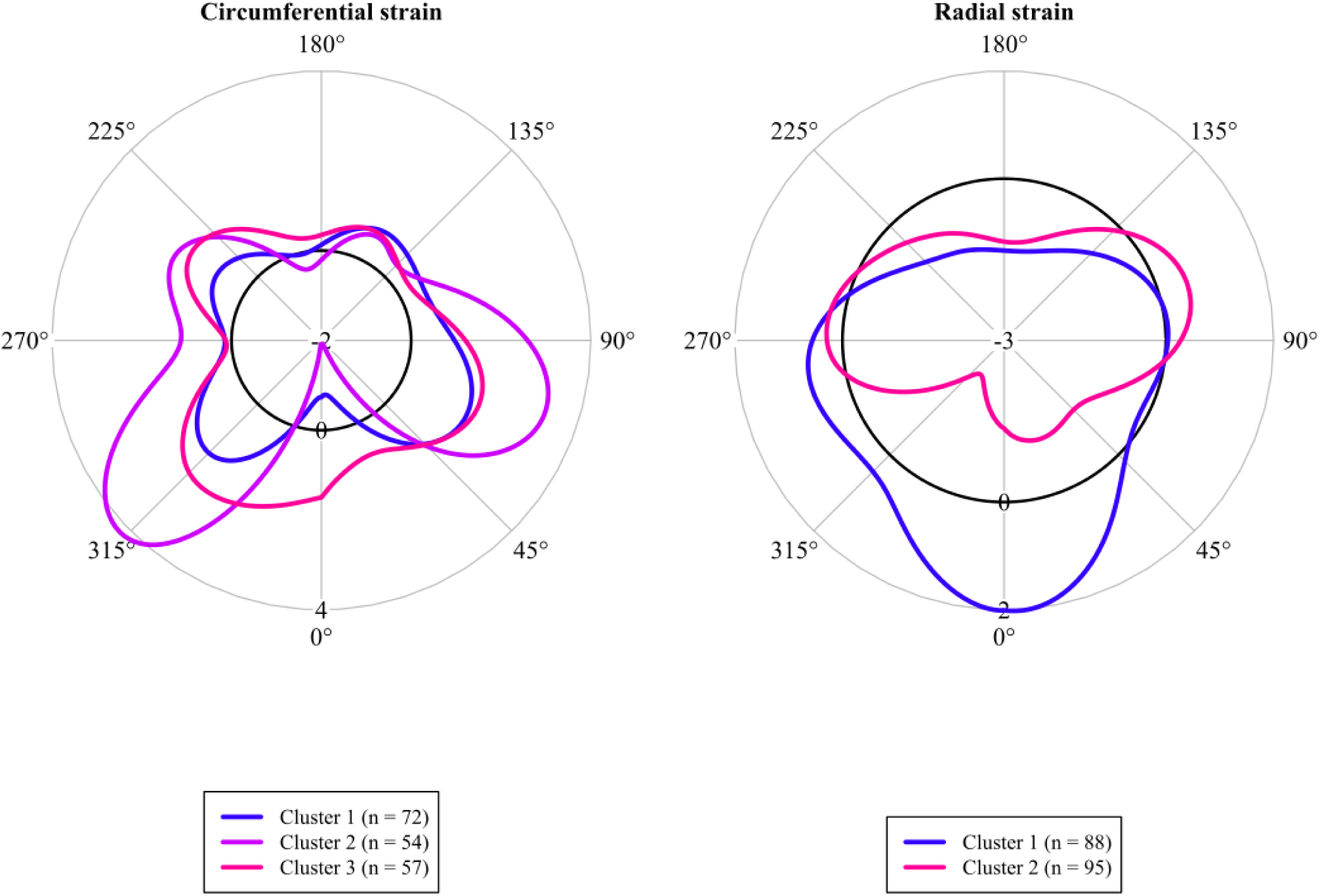
The mean circumferential and radial strain patterns each split into two clusters by the spectral consensus clustering algorithm.

**Figure S5:**
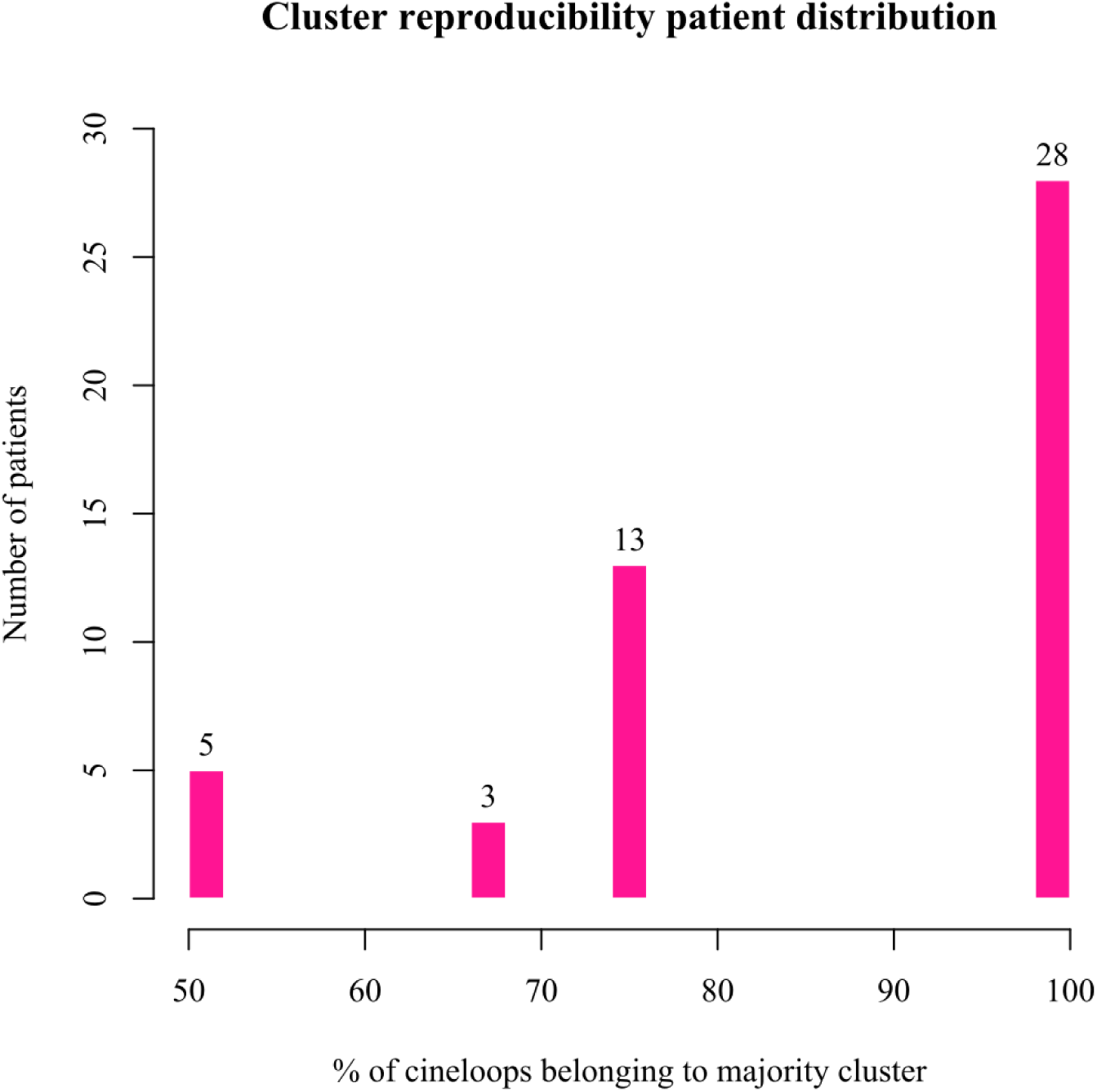
Distribution of patient cluster reproducibilities measured by the percentage of a patient’s cineloops belonging to the same cluster.

**Figure S6:**
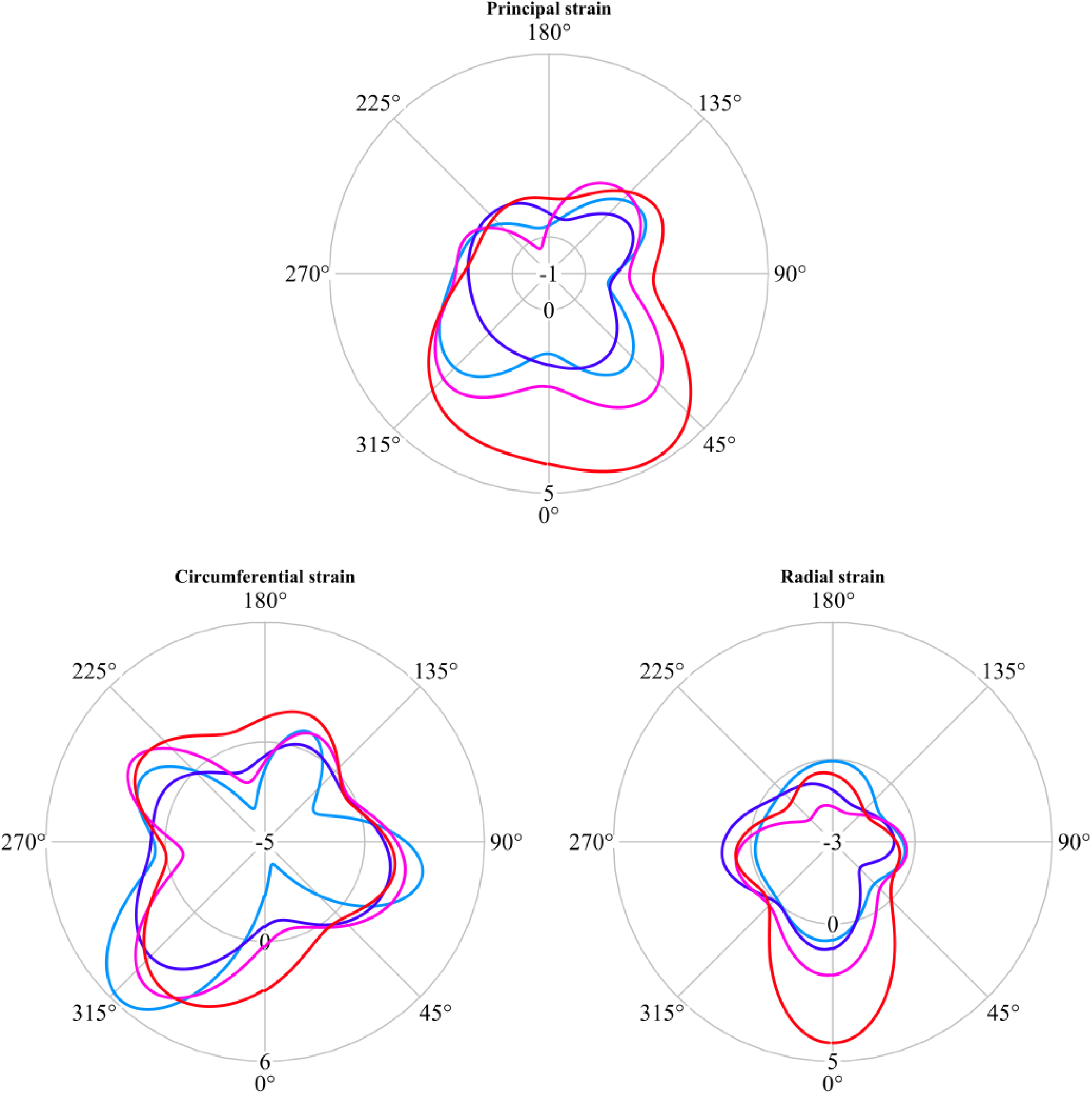
Strain patterns extracted from cineloops acquired on the same patients. Examples are chosen purely to illustrate how phases can vary little compared to the amplitude variation. Note, while each polar plot depicts strain patterns from the same patient, the principal, circumferential, and radial strain pattern polar plots are from different patients. These curves represent the smoothed strain patterns, before any alignment.

## REFERENCES

1. Lederle FA, Johnson GR, Wilson SE, Chute EP, Littooy FN, Bandyk D, et al. Prevalence and associations of abdominal aortic aneurysm detected through screening. Aneurysm Detection and Management (ADAM) Veterans Affairs Cooperative Study Group. Ann Intern Med. 1997;126(6):441–9.

2. Lindholt JS, Sogaard R. Population screening and intervention for vascular disease in Danish men (VIVA): a randomised controlled trial. Lancet. 2017;390(10109):2256–65.

3. Johansson G, Nydahl S, Olofsson P, Swedenborg J. Survival in patients with abdominal aortic aneurysms. Comparison between operative and nonoperative management. European journal of vascular surgery. 1990;4(5):497–502.

4. Karbase Årsrapport [Internet]. 2017.

5. Aggarwal S, Qamar A, Sharma V, Sharma A. Abdominal aortic aneurysm: A comprehensive review. Exp Clin Cardiol. 2011;16(1):11–5.

6. Umebayashi R, Uchida HA, Wada J. Abdominal aortic aneurysm in aged population. Aging (Albany NY). 2018;10(12):3650–1.

7. Wanhainen A, Verzini F, Van Herzeele I, Allaire E, Bown M, Cohnert T, et al. Editor’s Choice - European Society for Vascular Surgery (ESVS) 2019 Clinical Practice Guidelines on the Management of Abdominal Aorto-iliac Artery Aneurysms. Eur J Vasc Endovasc Surg. 2019;57(1):8–93.

8. Lederle FA, Wilson SE, Johnson GR, Reinke DB, Littooy FN, Acher CW, et al. Immediate repair compared with surveillance of small abdominal aortic aneurysms. N Engl J Med. 2002;346(19):1437–44.

9. UK Small Aneurysms Trial Participants. Mortality results for randomised controlled trial of early elective surgery or ultrasonographic surveillance for small abdominal aortic aneurysms. The UK Small Aneurysm Trial Participants. Lancet. 1998;352(9141):1649–55.

10. Filardo G, Powell JT, Martinez MA, Ballard DJ. Surgery for small asymptomatic abdominal aortic aneurysms. Cochrane Database Syst Rev. 2015;2015(2):CD001835.

11. Lederle FA, Johnson GR, Wilson SE, Ballard DJ, Jordan WD, Jr., Blebea J, et al. Rupture rate of large abdominal aortic aneurysms in patients refusing or unfit for elective repair. JAMA. 2002;287(22):2968–72.

12. Laine MT, Vänttinen T, Kantonen I, Halmesmäki K, Weselius EM, Laukontaus S, et al. Rupture of Abdominal Aortic Aneurysms in Patients Under Screening Age and Elective Repair Threshold. Eur J Vasc Endovasc Surg. 2016;51(4):511–6.

13. Lancaster EM, Gologorsky R, Hull MM, Okuhn S, Solomon MD, Avins AL, et al. The natural history of large abdominal aortic aneurysms in patients without timely repair. J Vasc Surg. 2022;75(1):109–17.

14. Nicholls SC, Gardner JB, Meissner MH, Johansen HK. Rupture in small abdominal aortic aneurysms. J Vasc Surg. 1998;28(5):884–8.

15. Kontopodis N, Pantidis D, Dedes A, Daskalakis N, Ioannou CV. The - Not So - Solid 5.5 cm Threshold for Abdominal Aortic Aneurysm Repair: Facts, Misinterpretations, and Future Directions. Front Surg. 2016;3:1.

16. Oliver-Williams C, Sweeting MJ, Jacomelli J, Summers L, Stevenson A, Lees T, et al. Safety of Men With Small and Medium Abdominal Aortic Aneurysms Under Surveillance in the NAAASP. Circulation. 2019;139(11):1371–80.

17. Hanna L, Borsky K, Abdullah AA, Sounderajah V, Marshall DC, Salciccioli JD, et al. Trends in Hospital Admissions, Operative Approaches, and Mortality Related to Abdominal Aortic Aneurysms in England Between 1998 and 2020. Eur J Vasc Endovasc Surg. 2023;66(1):68–76.

18. Howard DP, Banerjee A, Fairhead JF, Handa A, Silver LE, Rothwell PM, et al. Population-Based Study of Incidence of Acute Abdominal Aortic Aneurysms With Projected Impact of Screening Strategy. J Am Heart Assoc. 2015;4(8):e001926.

19. Long A, Rouet L, Bissery A, Rossignol P, Mouradian D, Sapoval M. Compliance of abdominal aortic aneurysms evaluated by tissue Doppler imaging: correlation with aneurysm size. Journal of vascular surgery. 2005;42(1):18–26.

20. Wilson KA, Lee AJ, Lee AJ, Hoskins PR, Fowkes FG, Ruckley CV, et al. The relationship between aortic wall distensibility and rupture of infrarenal abdominal aortic aneurysm. J Vasc Surg. 2003;37(1):112–7.

21. Wilson K, Bradbury A, Whyman M, Hoskins P, Lee A, Fowkes G, et al. Relationship between abdominal aortic aneurysm wall compliance and clinical outcome: a preliminary analysis. Eur J Vasc Endovasc Surg. 1998;15(6):472–7.

22. Sonesson B, Sandgren T, Lanne T. Abdominal aortic aneurysm wall mechanics and their relation to risk of rupture. Eur J Vasc Endovasc Surg. 1999;18(6):487–93.

23. Hoegh A, Lindholt JS. Vascular distensibility as a predictive tool in the management of small asymptomatic abdominal aortic aneurysms. Vasc Endovascular Surg. 2009;43(4):333–8.

24. Lorenzen US, Eiberg JP, Hultgren R, Wanhainen A, Langenskiold M, Sillesen HH, et al. The Short-term Predictive Value of Vessel Wall Stiffness on Abdominal Aortic Aneurysm Growth. Ann Vasc Surg. 2021;77:187–94.

25. Di Martino ES, Bohra A, Vande Geest JP, Gupta N, Makaroun MS, Vorp DA. Biomechanical properties of ruptured versus electively repaired abdominal aortic aneurysm wall tissue. J Vasc Surg. 2006;43(3):570–6; discussion 6.

26. Bracco MI, Broda M, Lorenzen US, Florkow MC, Somphone O, Avril S, et al. Fast strain mapping in abdominal aortic aneurysm wall reveals heterogeneous patterns. Front Physiol. 2023;14:1163204.

27. Zottola ZR, Kong DS, Medhekar AN, Frye LE, Hao SB, Gonring DW, et al. Intermediate pressure-normalized principal wall strain values are associated with increased abdominal aortic aneurysmal growth rates. Front Cardiovasc Med. 2023;10:1232844.

28. Derwich W, Keller T, Filmann N, Schmitz-Rixen T, Blase C, Oikonomou K, et al. Changes in Aortic Diameter and Wall Strain in Progressing Abdominal Aortic Aneurysms. J Ultrasound Med. 2023;42(8):1737–46.

29. Derwich W, Wittek A, Pfister K, Nelson K, Bereiter-Hahn J, Fritzen CP, et al. High Resolution Strain Analysis Comparing Aorta and Abdominal Aortic Aneurysm with Real Time Three Dimensional Speckle Tracking Ultrasound. Eur J Vasc Endovasc Surg. 2016;51(2):187–93.

30. Karatolios K, Wittek A, Nwe TH, Bihari P, Shelke A, Josef D, et al. Method for aortic wall strain measurement with three-dimensional ultrasound speckle tracking and fitted finite element analysis. Ann Thorac Surg. 2013;96(5):1664–71.

31. van Disseldorp EM, Petterson NJ, Rutten MC, van de Vosse FN, van Sambeek MR, Lopata RG. Patient Specific Wall Stress Analysis and Mechanical Characterization of Abdominal Aortic Aneurysms Using 4D Ultrasound. Eur J Vasc Endovasc Surg. 2016;52(5):635–42.

32. van Disseldorp EMJ, Petterson NJ, van de Vosse FN, van Sambeek M, Lopata RGP. Quantification of aortic stiffness and wall stress in healthy volunteers and abdominal aortic aneurysm patients using time-resolved 3D ultrasound: a comparison study. Eur Heart J Cardiovasc Imaging. 2019;20(2):185–91.

33. Broda M, Rouet L, Zielinski A, Sillesen H, Eiberg J, Ghulam Q. Profiling abdominal aortic aneurysm growth with three-dimensional ultrasound. Int Angiol. 2022;41(1):33–40.

34. Ullah S, Finch CF. Applications of functional data analysis: A systematic review. BMC Medical Research Methodology. 2013;13(1):43.

35. Mukaka MM. Statistics corner: A guide to appropriate use of correlation coefficient in medical research. Malawi Med J. 2012;24(3):69–71.

36. John CR, Watson D, Russ D, Goldmann K, Ehrenstein M, Pitzalis C, et al. M3C: Monte Carlo reference-based consensus clustering. Scientific Reports. 2020;10(1):1816.

37. John CR, Watson D, Barnes MR, Pitzalis C, Lewis MJ. Spectrum: fast density-aware spectral clustering for single and multi-omic data. Bioinformatics. 2020;36(4):1159–66.

38. Bracco MI, Eiberg JP, Lorenzen US, Avril S, Rouet L. Aortic Wall Stiffness Depends on Ultrasound Probe Pressure. 2023.

39. Svendsen MBS, Ghulam QM, Zielinski AH, Lachenmeier C, Eiberg JP. Validation of an assessment tool for estimation of abdominal aortic aneurysm compression in diagnostic ultrasound. Ultrasonics. 2021;116:106484.

40. Ghulam QM, Svendsen MBS, Zielinski AH, Eiberg JP. Ultrasound Transducer Pressure: An Unexplored Source of Abdominal Aortic Aneurysm Measurement Error. Ultrasound Med Biol. 2022;48(9):1778–84.

